# SAHVER: Subarachnoid Hemorrhage Volumetric Expediting Resolution

**DOI:** 10.1101/2024.09.09.24312803

**Authors:** Saif Salman, Yujia Wei, Melina Wirtz, Bradley J Erickson, Rohan Sharma, Nicholas Brandmeir, Behnam Rezai Jahromi, David Miller, Nadia Albaramony, Rabih Tawk, William David Freeman

## Abstract

**Introduction:** <u>***S***</u>ubarachnoid <u>***H***</u>emorrhage <u>***V***</u>olumetric <u>***A***</u>rtificial <u>***I***</u>ntelligence (<u>***SAHVAI***</u>) is a novel model that segments and quantifies Subarachnoid Hemorrhage Volume (SAHV) on non-contrast CT (NCCT) scans and generates a 3D brain volumetric map called SAHVAI-3D. It is enhanced into SAHVAI-4D when measured over time. Precise measurement of SAHV is critical to future discoveryies. For example, IRRAflow is a FDA-approved ventricular irrigation and drainage system that can expedite removal of SAH blood products.

**Objective:** Utilize SAHVAI model to compare and quantify the course of SAHV resolution over time and generate SAHVAI-3D brain maps to help visualize significant SAHV resolution patterns and predict vasospasm.

**Methods:** We applied SAHVAI to SAH cases with mFS(3-4) using the NCCT scans among three groups. Group A included 1 SAH patient treated with the IRRAflow system. Group B included one SAH patient presented GCS 15 two days after ictus with no requirement for EVD. Group C included 10 patients who underwent regular EVD placement per standard of care.

**Results:** Group A showed expedited resolution of SAHV (1.87mL/day) with an mRS of 0 on discharge and minimal vasospasm (Figures 1, 2). Group B showed 16mL increase in SAHV suspected for aneurysmal rebleeding days (5-9), and the patient later died (mRS of 6) (Figure 3). Group C showed reduction of SAHV of ∼ 0.5ml /day (Figure 4). Further, the resultant 3D brain maps revealed that areas with the highest density of blood concentration were correlated with the severity and location of the vasospasm in all groups (Table 1).

**Conclusion:** SAHVAI, SAHVAI-3D and SAHVAI-4D are novel methods that reliably quantifies SAHV blood volume and changes over time including SAH blood resolution or rebleeding events. SAHVER is a model that shows how interventions such as IRRAflow can expedite SAHV resolution compared to passive EVD and non-CSF drainage groups.

## Introduction

Subarachnoid hemorrhage (SAH) is a critical medical condition that involves bleeding into the subarachnoid space, often due to ruptured aneurysms. Accurate measurement and timely management of SAH are crucial for improving patient outcomes. Traditional methods for assessing SAH have relied on clinical scales and qualitative imaging assessments. However, these methods may lack precision and consistency, particularly in quantifying hemorrhage volume and its resolution over time^1-10^.

The introduction of Subarachnoid Hemorrhage Volumetric Artificial Intelligence (SAHVAI) represents a significant advancement in the quantification and visualization of SAH. The SAHVAI model is capable of segmenting and quantifying SAHV on non-contrast CT (NCCT) scans, generating a 3D brain volumetric map, referred to as SAHVAI-3D. When this model is applied to serial scans over time, it evolves into SAHVAI-4D, offering a dynamic view of SAH resolution or progression^11-15^.

Precise measurement of SAHV is not just an academic exercise; it has practical implications for patient management and outcome prediction. For instance, the IRRAflow system, an FDA-approved device for ventricular irrigation and drainage, has shown promise in expediting the removal of SAH blood products, potentially leading to better outcomes^16-18^.

This study aims to utilize the SAHVAI model to compare and quantify the course of SAH resolution over time across different patient groups. By generating SAHVAI-3D brain maps, the study seeks to visualize significant patterns in SAH resolution and predict vasospasm, a common and severe complication of SAH.

## Methods

### Study Design

This study was conducted retrospectively, analyzing SAH cases with a modified Fisher Scale (mFS) of 3-4. The mFS is a commonly used grading system that quantifies the amount of subarachnoid and intraventricular blood seen on CT scans and predicts the risk of vasospasm.

### Patient Groups

The study included three distinct patient groups:

- **Group A:** One SAH patient treated with the IRRAflow system, designed for ventricular irrigation and drainage. This patient was selected to examine the potential benefits of active SAH blood removal in comparison to other methods.
- **Group B:** One SAH patient who rebleed on day 5 but initially presented with a Glasgow Coma Scale (GCS) score of 15 two days post-ictus and did not require EVD. This case was included to represent a patient with minimal clinical intervention.
- **Group C:** Ten SAH patients who underwent standard EVD placement per standard of care. This group was included to serve as a comparison to the IRRAflow-treated patient and to examine the outcomes with conventional treatment methods.

### SAHVAI Implementation

The SAHVAI model was applied to NCCT scans of all patients across the three groups. The model automatically segmented and quantified SAHV, generating 3D brain maps (SAHVAI-3D). When applicable, serial scans were analyzed to produce SAHVAI-4D maps, providing insights into the dynamic changes in SAHV over time.

### Outcome Measures

The primary outcome measure was the rate of SAH resolution, quantified as the reduction in SAHV per day. Secondary outcome measures included the modified Rankin Scale (mRS) at discharge, incidence of vasospasm, and the correlation between blood concentration density in specific brain regions and vasospasm severity.

## Results

### Group A (IRRAflow Treated)

The SAH patient treated with the IRRAflow system demonstrated an expedited resolution of SAHV, with an average reduction of 1.87 mL per day. This patient had a mRS of 0 at discharge, indicating no significant disability, and experienced minimal vasospasm. The SAHVAI-3D map for this patient revealed a uniform and rapid decrease in SAH volume across all brain regions. (figure1, 2)

**Figure 1.**
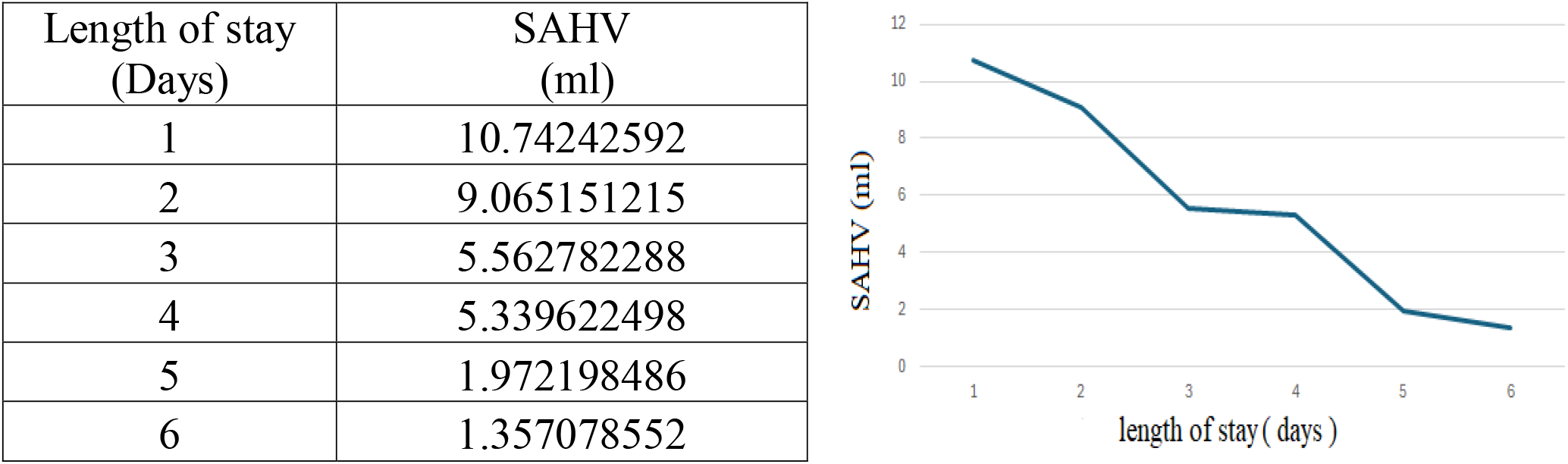
The quantified SAHV measured via SAHVAI shows expedited resolution of SAHV (1.87mL/day) when using the active Irrigation and drainage system (the IRRAFlow).

**Figure 2.**
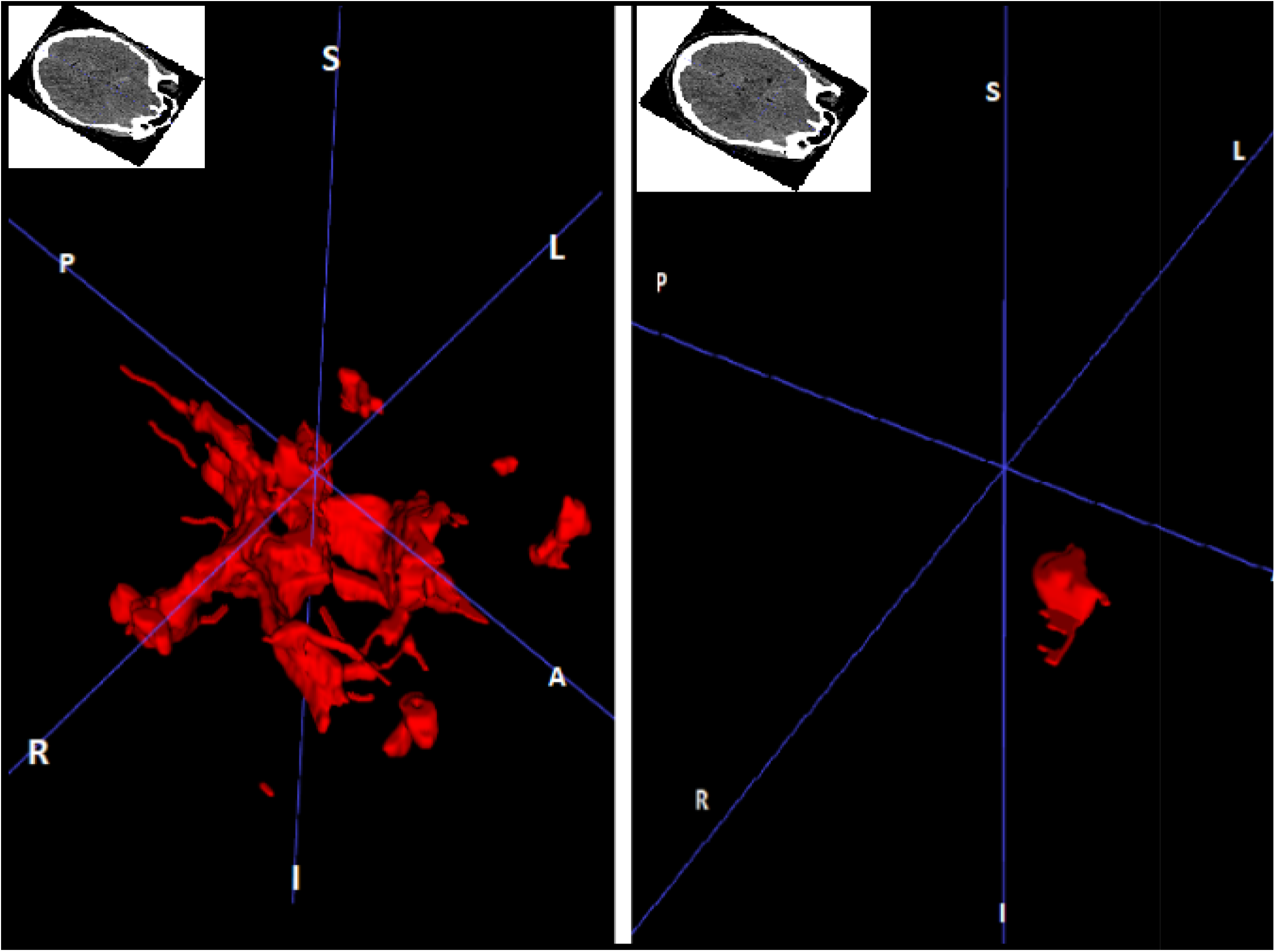
SAHVAI-3D brain maps showing the distribution of SAH before versus after the active irrigation and drainage. (A=Anterior, P= Posterior, L= Left, R=Right, S=superior, I=Inferior).

### Group B (rebleed)

In contrast, the patient in Group B, who did not require EVD, exhibited a 16 mL increase in SAHV between days 5 and 9, suggestive of aneurysmal rebleeding. Unfortunately, this patient had a poor outcome, with an mRS of 6 (death). The SAHVAI-3D map showed an initial stable SAHV, followed by a significant increase in volume localized to the region of rebleeding.(figure 3)

**Figure 3.**
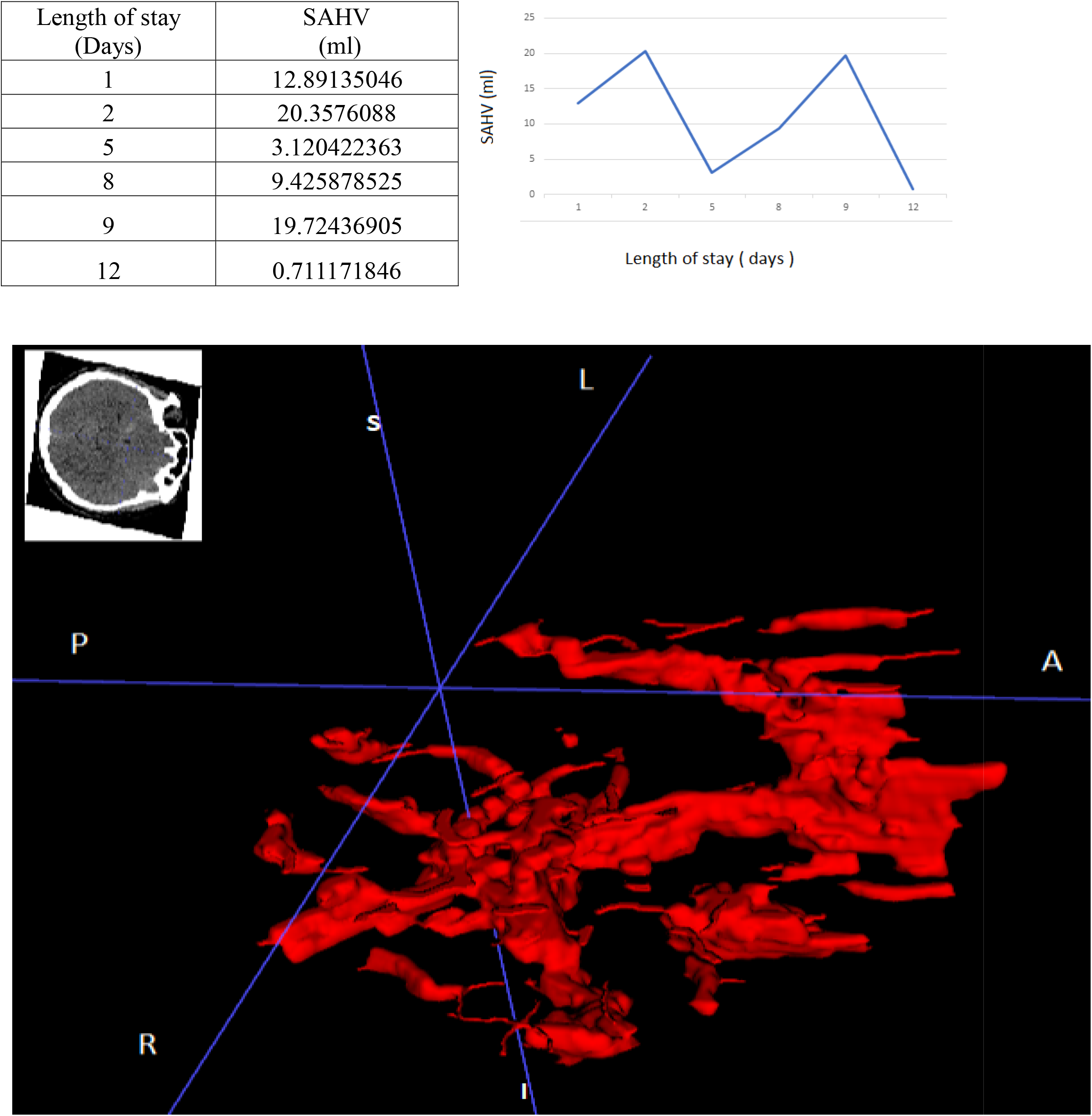
(a)The quantified SAHV measured via SAHVAI showing the rebleeding on day 9. (b) SAHVAI-3D brain map showing the rebleed on days (5-9) in the basal cistern, midline-frontal, and ACA distribution. (A=Anterior, P= Posterior, L= Left, R=Right, S=superior, I=Inferior).

### Group C (Standard EVD Treated)

Patients in Group C, who received standard EVD treatment, showed a more gradual reduction in SAHV, averaging approximately 0.5 mL per day. The mRS at discharge for this group varied, with some patients achieving good recovery (mRS 1-2) and others having moderate to severe disability (mRS 3-5). The SAHVAI-3D maps for these patients highlighted areas of persistent blood concentration, which correlated with the severity and location of vasospasm observed clinically (figure 4), (table 1).

**Figure 4.**
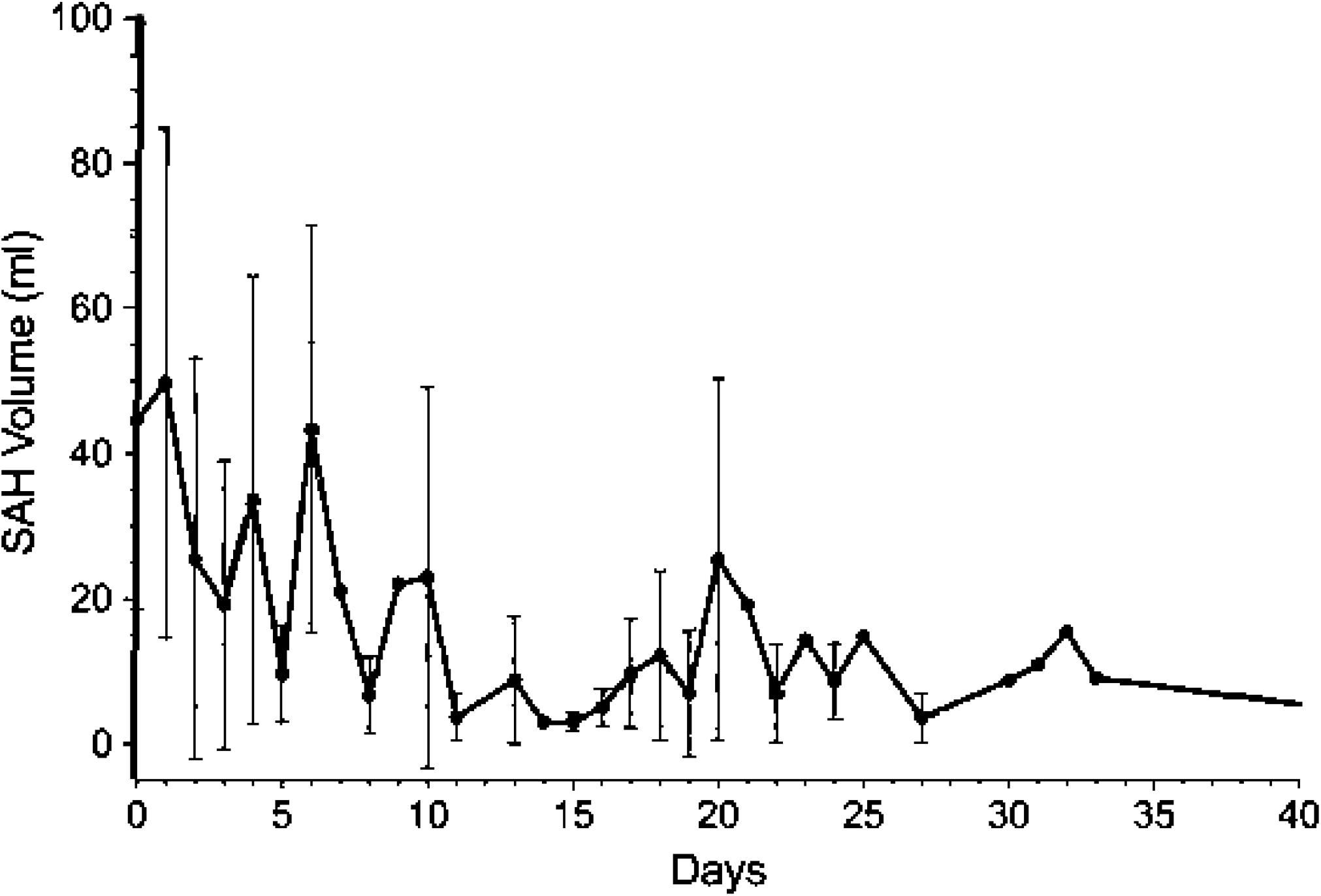
Plot of the Mean and Standard Deviation measured the quantitative SAH volume (SAHV) over time of the total cohort (n=10) day by day, showed reduction of SAHV of ∼ 0.5ml /day

**Table 1.** the correlation between areas showing the highest blood concentration density with the vasospasm’s severity and location.

|  | Case | Severity of Vasospasm | Location of VSP if obtainable | Visual area of thickest blood concentration on Brain Map |
| --- | --- | --- | --- | --- |
| Group A | 1 | minimal | / | / |
| Group B | 1 | severe | Bilateral ACA | midline-frontal, ACA distribution |
| Group C | 1 | severe | right MCA | right convexity of the brain |
|  | 2 | / | / | diffuse |
|  | 3 | mild | right MCA | right convexity of the brain |
|  | 4 | severe | right MCA | right convexity of the brain |
|  | 5 | severe | diffuse | posterior and right convexity of the brain |
|  | 6 | severe | bilateral ACA | left convexity of the brain (upper part of the brain) |
|  | 7 | moderate - severe | diffuse | left convexity of the brain |
|  | 8 | mild | right VA | posterior location of the brain (brainstem) |
|  | 9 | severe | bilateral MCA, ACA, diffuse | diffuse, especially right convexity of the brain |

### SAHVAI-3D and 4D Maps

The 3D and 4D brain maps generated using SAHVAI provided valuable insights into the spatial and temporal dynamics of SAH resolution. The areas with the highest blood concentration density were found to correlate strongly with the severity of vasospasm across all groups. These findings suggest that SAHVAI-3D and 4D maps could be used as predictive tools for vasospasm, allowing for earlier and more targeted interventions.

## Discussion

The results of this study underscore the potential of the SAHVAI model in improving the management of SAH. By providing precise and consistent quantification of SAHV, SAHVAI enables clinicians to monitor the effectiveness of treatments like the IRRAflow system and standard EVD. The expedited SAH resolution observed in the IRRAflow-treated patient compared to those receiving standard EVD suggests that active removal of blood products may lead to better outcomes.

Furthermore, the ability of SAHVAI to generate 3D and 4D maps of the brain offers a novel approach to visualizing SAH resolution and predicting complications like vasospasm. The correlation between high blood concentration density and vasospasm severity observed in this study highlights the potential of these maps as predictive tools, which could guide clinical decision-making and improve patient outcomes^19-22^.

## Conclusion

SAHVAI, along with its 3D and 4D mapping capabilities, represents a significant advancement in the quantification and visualization of SAH. This model not only provides a more accurate assessment of SAH volume but also offers the potential to predict and prevent complications such as vasospasm. The findings from this study suggest that interventions like the IRRAflow system may expedite SAH resolution compared to standard treatment methods, leading to better clinical outcomes. Future studies should focus on validating these findings in larger patient cohorts and exploring the use of SAHVAI in other types of intracranial hemorrhage.

## Data Availability

All data produced in the present study are available upon reasonable request to the authors

## Notes

### Competing Interest Statement

The authors have declared no competing interest.

### Funding Statement

This study did not receive any funding

### Author Declarations

Ethics committee/IRB of The Mayo Clinic waived ethical approval for this work

